# Network biology and bioinformatics-based framework to identify the impacts of SARS-CoV-2 infections on lung cancer and tuberculosis

**DOI:** 10.1101/2024.09.10.24313452

**Authors:** Abdul Waaje, Md Sumon Sarkar, Md Zahidul Islam

## Abstract

The severe acute respiratory syndrome coronavirus 2 (SARS-CoV-2) is a coronavirus variation responsible for COVID-19, the respiratory disease that triggered the COVID-19 pandemic. The primary aim of our study is to elucidate the complex network of interactions between SARS-CoV-2, tuberculosis, and lung cancer employing a bioinformatics and network biology approach. Lung cancer is the leading cause of significant illness and death connected to cancer worldwide. Tuberculosis (TB) is a prevalent medical condition induced by the Mycobacterium bacteria. It mostly affects the lungs but may also have an influence on other areas of the body. Coronavirus disease (COVID-19) causes a risk of respiratory complications between lung cancer and tuberculosis. SARS-CoV-2 impacts the lower respiratory system and causes severe pneumonia, which can significantly increase the mortality risk in individuals with lung cancer. We conducted transcriptome analysis to determine molecular biomarkers and common pathways in lung cancer, TB, and COVID-19, which provide understanding into the association of SARS-CoV-2 to lung cancer and tuberculosis. Based on the compatible RNA-seq data, our research employed GREIN and NCBI’s Gene Expression Omnibus (GEO) to perform differential gene expression analysis. Our study exploited three RNA-seq datasets from the Gene Expression Omnibus (GEO)—GSE171110, GSE89403, and GSE81089—to identify distinct relationships between differentially expressed genes (DEGs) in SARS-CoV-2, tuberculosis, and lung cancer. We identified 30 common genes among SARS-CoV-2, tuberculosis, and lung cancer (25 upregulated genes and 5 downregulated genes). We analyzed the following five databases: WikiPathway, KEGG, Bio Carta, Elsevier Pathway and Reactome. Using Cytohubba’s MCC and Degree methods, We determined the top 15 hub genes resulting from the PPI interaction. These hub genes can serve as potential biomarkers, leading to novel treatment strategies for disorders under investigation. Transcription factors (TFs) and microRNAs (miRNAs) were identified as the molecules that control the differentially expressed genes (DEGs) of interest, either during transcription or after transcription. We identified 35 prospective therapeutic compounds that form significant differentially expressed genes (DEGs) in SARS-CoV-2, lung cancer, and tuberculosis, which could potentially serve as medications. We hypothesized that the potential medications that emerged from this investigation may have therapeutic benefits.

## 1. Introduction

A global pandemic of respiratory tract infections known as “coronavirus disease 2019” (COVID-19) was caused by the extremely contagious and lethal coronavirus named severe acute respiratory syndrome coronavirus 2 (SARS-CoV-2which first manifested itself in December 2019 [1]. According to the World Health Organization (WHO), there have been 771 million confirmed COVID-19 cases worldwide, including 6 million fatalities by the end of 2023. SARS-CoV-2 is liable for inducing a severe form of pneumonia that poses a significant risk to human life. This pneumonia is characterized by the death of alveolar cells or the occurrence of diffuse alveolar damage (DAD), which has the potential to progress into acute respiratory distress syndrome (ARDS) [2]. Tuberculosis (TB) is a prevalent health condition caused by the Mycobacterium bacteria, primarily impacting the pulmonary system but occasionally affecting various extrapulmonary regions of the body [3]. The transmission of tuberculosis-causing bacteria occurs via airborne routes, primarily through inhalation or exposure to respiratory droplets containing the pathogen. The bacteria colonize the pulmonary region before disseminating via the bloodstream to many anatomical sites, including but not limited to the heart, kidneys, and other organs [4]. The World Health Organization (WHO) predicts that approximately 10.6 million individuals will be diagnosed with tuberculosis (TB) in 2021, and approximately 1.6 million individuals will fall victim to the active form of the disease [5]. Consequently, TB stands as one of the foremost contributors to global mortality rates. Globally, lung cancer is the most prevalent cause of cancer-related illness and death [6]. Smoking tobacco is well recognized as the primary factor contributing to the development of lung cancers, with a substantial majority of cases (ranging from 80% to 90%) occurring in individuals who smoke cigarettes. Additional professional and external variables that are frequently associated with the development of lung cancer encompass exposure to metal dust and radiation [7]. According to data provided by the International Agency for Research on Cancer and the World Health Organization in 2020, Lung cancer (LC) is the most elevated fatality rate compared to other forms of cancer.

Approximately 1.79 million deaths were attributed to LC, representing 18% of all malignancies. This percentage is considerably greater than that observed for other forms of cancer. The cumulative survival rate for lung cancer over a period of 5 years is below 20% [8]. Several researches have emphasized the potential escalation of tuberculosis incidence and mortality rates amid the COVID-19 pandemic [9][10]. This assertion is substantiated by the World Health Organization’s data from October 2021, which indicates a rise in global tuberculosis-related fatalities for the first time in ten years, attributable to the effect of the COVID-19 pandemic [11]. Globally, the COVID-19 pandemic exacerbated the tuberculosis (TB) epidemic due to dispersion of treatment facilities and additional demands on medical facilities, leading to a reduction of national TB applications [12]. Recent research has linked TB to increased fatality in COVID-19 patients. Coronavirus disease (COVID-19) causes a risk of respiratory complications among individuals with lung cancerSARS-CoV-2 primarily targets the lower respiratory tract and leads to life-threatening pneumonia, which can contribute to an elevated mortality rate among lung cancer sufferers [13]. Patients with lung cancer have a greater propensity for severe COVID-19 complications and greater mortality rates (33–42%) than individuals with other tumors [14]. Numerous studies have shown that pulmonary TB is associated with a higher probability and morbidity of lung cancer, and vice versa [15]. In countries with high tuberculosis stress, the prevalence of tuberculosis among people with cancer was as elevated as 12.72 percent. Several theories suggest that infection with M. tuberculosis may lead to the development of lung cancer [16]. These hypotheses encompass the suppression of the immune system, DNA damage, and the generation of autoimmune factors. The fundamental proposition posits that *Mycobacterium tuberculosis* is responsible for inducing persistent inflammation, potentially resulting in the development of lung carcinoma [17].

Modern technologies are necessary to manage challenging situations in human existence. X-ray, CT, and ultrasound image processing and analysis can be utilized to diagnose COVID-19 using machine learning (ML) and artificial intelligence (AI). These techniques can be applied to distinguish COVID-19 from other pneumonia causes. Employing techniques such as, support vector machines,neural networks, decision trees, and models for machine learning to predict both mortality and severity, AI can forecast, diagnose, and model COVID-19 [18]. However, a number of innovative technologies are available to provide COVID-19 patients with healthcare services. These techniques included social media, telemonitoring, teleradiology, telesurgery, cloud-based services, telemedicine, tele-consulting, video conferences, Virtual visits, virtual healthcare, e-consultations, mobile-based self-care, and tele-ICU [19]. Utilizing telemedicine and other technology improves access to timely, high-quality care while preserving physical distance, reducing illness, and providing home COVID-19 inspection and monitoring during epidemics. Telemedicine has the potential to provide services without the necessity of face-to-face contact and the time required to obtain a diagnosis and commence treatment [20].

Numerous substantial empirical findings suggesting that SARS-CoV-2, tuberculosis, and lung cancer are clinically and histologically significant. Nevertheless, these complex relationships have hitherto remained unexplored. The primary goal of our study is to uncover the intricate structure of connections among SARS-CoV-2, tuberculosis & lung cancer by employing a bioinformatics and network biology methodology. With this undertaking, our aim is to offer a comprehensive understanding of how SARS-CoV-2 infections impact these two distinct but interconnected medical problems. Three datasets were utilized in this inquiry to clarify the biological correlations among SARS-CoV-2, tuberculosis, and lung cancer. The Gene Expression Omnibus (GEO) database was queried for these datasets, specifically GSE171110, GSE89403, and GSE81089, which correspond to SARS-CoV-2, tuberculosis, and lung cancer, respectively. Differentially expressed genes (DEGs) were determined for each dataset. Following this, DEGs that were shared among the three diseases were identified. The experimental genes that comprise the primary set for our entire investigation are these DEGs that are shared. By making use of these common differential regions (DEGs), we performed additional experiments and analyses, including enrichment and pathway analysis to elucidate the biological procedure underlying Genome-based expression analyses. The identification of hub genes, which are primarily concentrated in the common DEGs is a critical element in the prediction of potential medications. In order to aggregate the hub genes, we also established a network of protein-protein interactions (PPIs) using the shared DEGs. Furthermore, transcriptional regulators were identified by utilizing the shared DEGs of datasets. In conclusion, we put forward prospective therapeutic candidates. The simplified representation of the methodology that underpins this study is depicted in Figure 1.

**Figure 1:**
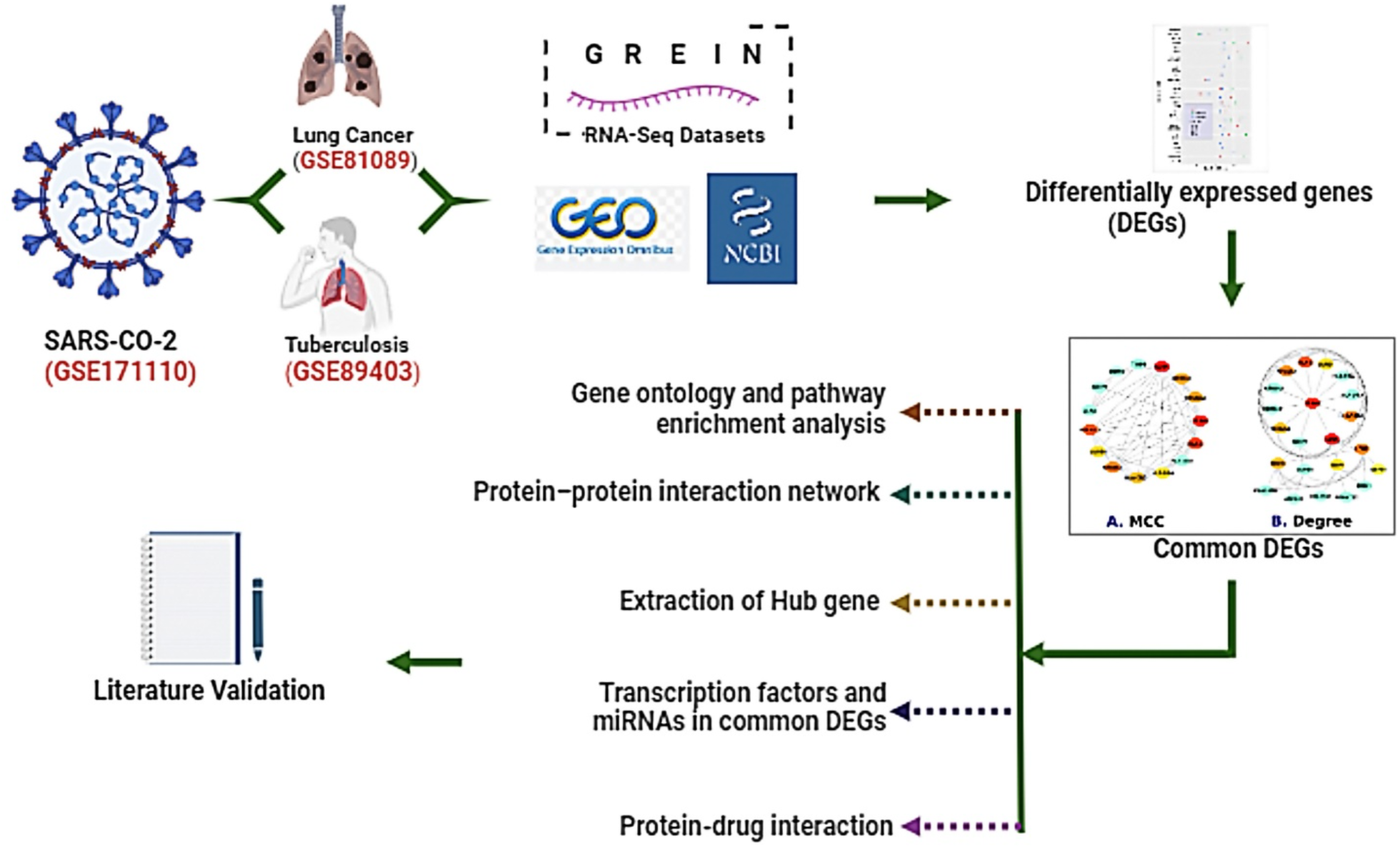
Illustrative outline of the methodology of this investigation.

## 2. Materials and Methods

### 2.1 Datasets utilized in this investigation

We looked at data from GREIN and the NCBI’s Gene Expression Omnibus (GEO) [21]. A website-based interactive application called GREIN allows for easy access to GEO RNA-seq data. Several studies used GREIN to perform differential gene expression analysis on the relevant RNA-seq data [22]. Less than six samples, a lack of control and case requirements, repeated datasets, unfavorable formatting, a concentration on unrelated experiments, or non-human RNAseq data were among the reasons why several datasets were rejected. A minimum of three case samples and one control sample must be included in each of the six samples. We looked for a dataset with a sufficient number of controls and cases for our study. Utilizing this method, we were able to identify three datasets that were relevant to our investigation and related to lung cancer, tuberculosis, and COVID-19. For our study, the genomic expression datasets GSE81089 [23], GSE89403 [24], and GSE171110 [25] were examined, each with a healthy control group and a case group. The NSCLC dataset (GSE81089) from Homo sapiens was used to compare the RNAseq data of 199 NSCLC samples. The tuberculosis dataset (GSE89403) used blood RNA indicators to forecast how patients would respond to treatment and healthy control group. GSE171110 was blood RNA-seq gene expression analysis in severely hospitalized patients compared to healthy donors.

**Figure 2:**
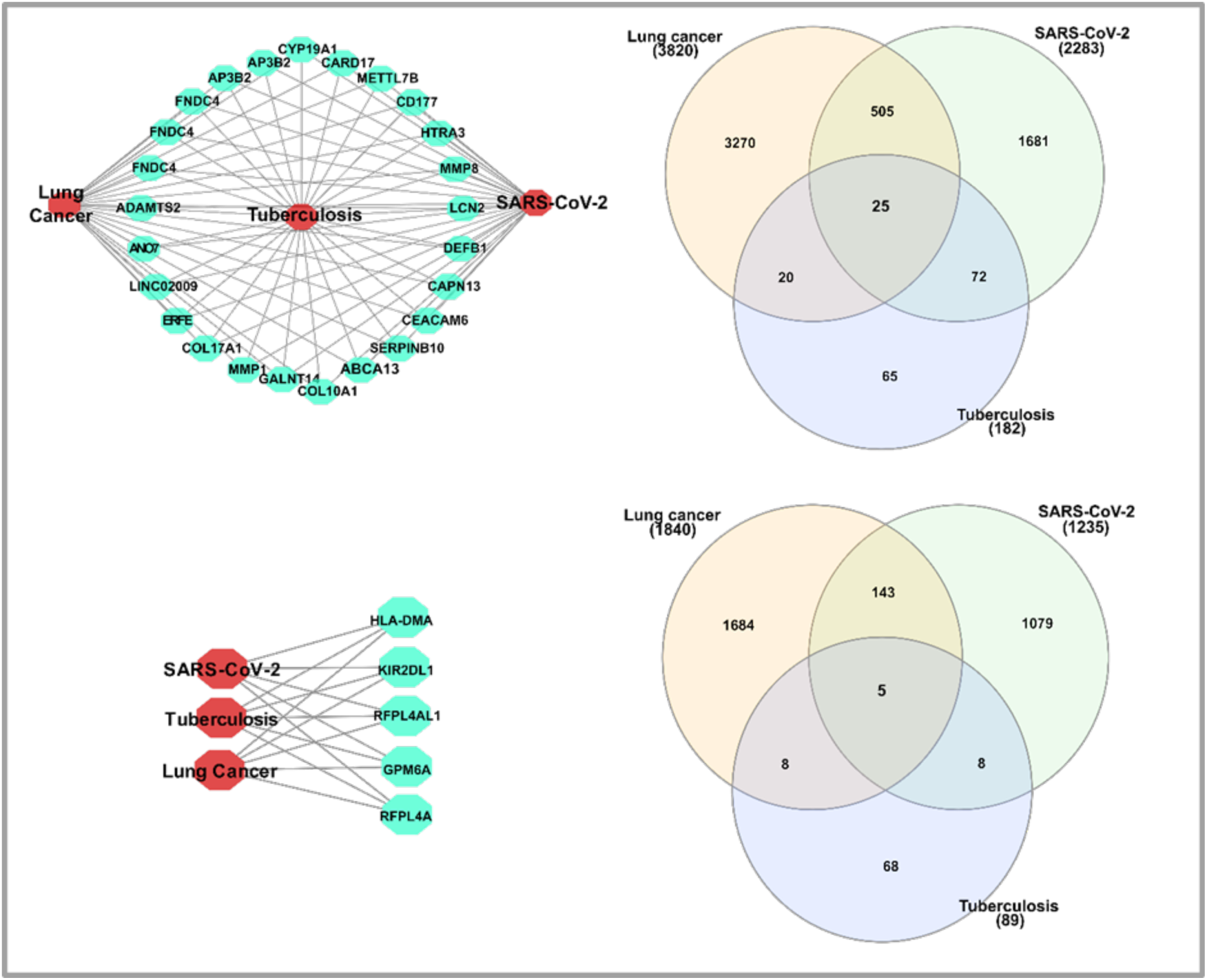
The differential representation of upregulated and downregulated genes is observed in the contexts of SARS-CoV-2, lung cancer, and tuberculosis.

### 2.2 Pre-processing of the raw data and analysis of differential expression

Differentially expressed genes (DEGs) can function as biomarkers, providing evidence of the existence or advancement of a disease [26]. The gene expression microarray datasets utilized in our study were made available by the NCBI Gene Expression Omnibus. By comparing healthy tissue with diseased tissue, the datasets were generated by finding differentially expressed genes (DEGs) connected to all pathology. Obtaining DEGs from the datasets GSE81089, GSE89403, and GSE171110 was the main goal of this investigation. We determined the range of Identification of Differentially Expressed Genes (DEGs) using two distinct criteria: absolute log fold change, abs (LogFC) >= +1.0 and adjusted p-values<0.05 for upregulated genes. Furthermore, downregulated genes exhibited, absolute log fold change, abs (LogFC)<= −1.0 and adjusted p-values<0.05. The mutual DEGs of GSE81089, GSE89403, and GSE171110 were then obtained using the VENN analysis Jvenn [27].

**Figure 3:**
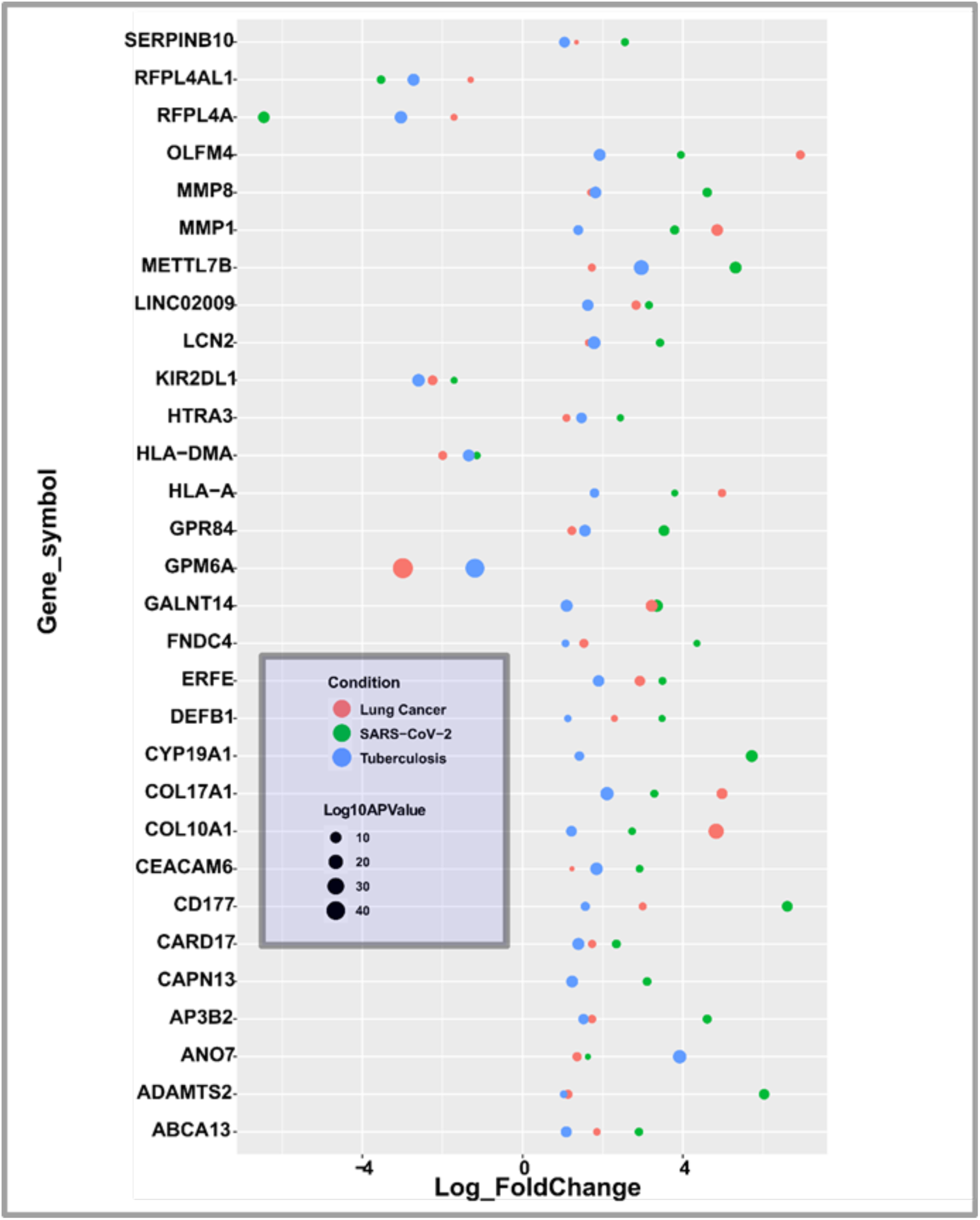
An outline of the transcriptome analysis is presented in the diagram. A. Determine the shared indicator genes between SARS COV-2, lung cancer, and tuberculosis by employing a Venn diagram. B. Common gene bubble graph using log10APValue and Log2 fold change. Log2 fold change shows gene expression variation, and log10APValue shows gene significance.

### 2.3 Gene ontology and pathway enrichment analysis

Gene ontology and pathway enrichment analyses are utilized to ascertain the involvement of significant genes, pathways, and biological processes [28]. In order to elucidate signal transduction pathway and the molecular mechanisms of commonly differentially expressed genes (DEGs), researchers conducted studies using EnrichR, a comprehensive web tool for complete gene arrays. These studies focused on gene ontology, functional enrichment (including biological procedures, cellular components, and molecular functions), and pathway enrichment [29]. By focusing on the parts and purposes of genes, gene ontology offers vast computational information resources. An ontological idea describes a body of knowledge within a particular context [30]. At this point, we examined the following five databases: Elsevier Pathway [32] Bio Carta [31], WikiPathway [34], KEGG [32], Reactome [33], and KEGG [32], These genomic data repositories seek to offer a comprehensive view of the molecular connections and metabolic pathways and activities of genes in many animals. These pathways were then separated into functional groups for examination. On the other hand, the GO approach divided them into operational groups using the terms biological process (BP), molecular function (MF), cellular component (CC) [35].

### 2.4. Analysis of networks involving protein–protein interactions

Protein-protein interactions (PPIs) are crucial for understanding cellular functions and diseases, playing a key role in drug discovery and therapeutic development. Understanding and obtaining insights into the activities of intracellular components is primarily and fundamentally achieved through the examination and Investigation of the PPI structure and functions [36][37][38]. Using the STRING (version 11.5) library, we developed the PPI connection obtained from shared DEGs to illustrate the Operational and physical links between lung cancer, TB, and COVID-19 [39]. Using topological characteristics from PPI analysis (degree above 15°), highly interacting proteins were discovered. The goal of STRING is to offer deeper understandings of PPI refers to the measurement of category confidence ratings across multiple levels: low, moderate, and high. To facilitate graphical analysis and conduct more PPI connection investigations, we have uploaded our PPI network in Cytoscape (version 3.7.0) [40]. The widely used network representation platform Cytoscape (v.3.7.0) combines numerous datasets to provide superior results for a variety of interactions, including PPIs, DNA-protein couplings, DNA interconnections, and more.

### 2.5. Detection of hub genes and analysis of submodules

Hub genes are essential for identifying significant molecular interactions and pathways that may influence disease development and therapy responses [41]. These genes have a sizable impact on the characteristics and development of the disease [42]. We used the Cytohubba Cytoscape plugin to order and identify important or potential target biological network components based on several network features. Cytohubba offers 11 methods for studying networks from various angles [43]. We identified the most significant hub genes inside the PPI relationship using Cytohubba’s MCC and Degree approach.

### 2.6. Participation of microRNAs and transcription factor in common DEGs

The dysregulation or abnormal activity of transcription factors are crucial for regulating gene expression, may provide the pathogenesis of certain diseases [44] Essential proteins called transcription factors (TFs) interpret the genetic code, enabling every kind of cell in our body to produce a distinct set of RNA and protein molecules. Essential proteins called transcription factors (TFs) interpret the genetic code, enabling every kind of cell in our body to produce a distinct set of RNA and protein molecules [45]. To identify the regulating molecules (i.e., miRNAs and TFs) that regulate DEGs of significance during or after transcription, we looked at the interactions between transcription factors (TFs) and DEGs as well as between microRNAs and DEGs. We used the NetworkAnalyst system to find TFs that are trying to link to our shared DEGs in the Chip-X [47] JASPAR [46] databases. The two main databases with experimental validity for genes that miRNAs target are Tarbase [48] and MirTarbase [49]. According to the degree above or equivalent to 20 from the TFs -DEGs connection, the TFs were sorted. We chose the miRNAs based on the DEGs-miRNA network degree above or equal to 15. The topological investigation made use of Network Analyst [50] and Cytoscape’s [51]. Using Cytoscape, researchers can choose the most important miRNAs with significant degrees, pinpoint biological functions and traits, and offer a workable biological hypothesis.

## 3. Results

### 3.1 Evaluation of COVID-19, lung cancer, and TB differential expression genes

The statistical data for SARS-CoV-2, lung cancer, and tuberculosis, are presented in the Table 1. Table 1 contains the following data: disease name, cellular features, GEO identification number, data type, total number of genes for each dataset, number of control and case specimens, significant genes, upregulated genes, significant genes and downregulated genes. Differential expression analysis, (also referred to as signature data collection) identified a total of 21180 genes associated with SARS-CoV-2. We considered Adjusted P-Value and “Log2 Fold-Change” when evaluating most significant genes. We determined 2283 up-regulated genes and 1235 down-regulated genes in SARS-CoV-2. The tuberculosis dataset contained 138 samples in total (38 for the control) and (100 for the case 2321) genes in signature data. Then, employing the same criteria, 271 significant genes were found (were up-regulated genes, and 89 were down-regulated). Again, the dataset for lung cancer included 218 samples in total (19 for the control and 199 for the case) and 25139 genes were identified after the generation of signature data. Then employing the same criteria again 5660 significant genes are identified (3820 were up-regulated genes, and 1840 were down-regulated genes). Next, a comparative analysis was performed to examine the up-regulated genes that are linked to tuberculosis, SARS-CoV-2, and lung cancer. We also compared the down-regulated genes across all conditions. We isolated 25 frequent up-regulated genes and 5 frequent down-regulated genes among SARS-CoV-2, tuberculosis, and lung cancer. The most significant frequent up-regulated genes among SARS CoV-2, tuberculosis and lung cancer were COL10A1, GALNT14, MMP1, COL17A1, ERFE, LINC02009, ANO7, ADAMTS2, FNDC4, OLFM4, GPR84, AP3B2, HLA-A, CYP19A1, CARD17, METTL7B, CD177, HTRA3, MMP8, LCN2, DEFB1, CAPN13, CEACAM6, and SERPINB10. The most significant frequent down-regulated genes among SARS-CoV-2, tuberculosis, and lung cancer were GPM6A, KIR2DL1, HLA-DMA, RFPL4A, and RFPL4AL1.

**Table 1:** The analysis and summary of the transcriptomic data. The information contained within it includes the identification code of the dataset, the sampling location, the precise quantity of the raw gene, the value of the specimen, and the significant genes.

| Disease Name | Tissue | GSE Number | Data Type | RAW genes | Number of Control and Case Sample | Total | Up | Down |
| --- | --- | --- | --- | --- | --- | --- | --- | --- |
| SARS-CoV-2 | Blood | GSE171110 | RNA Grein | 21180 | Case = 44<br>Control = 10 | 3518 | 2283 | 1235 |
| Lung Cancer | Non-small cell lung cancer (NSCLC) Tissue | GSE81089 | RNA Grein | 25139 | Case = 199<br>Control = 19 | 5660 | 3820 | 1840 |
| Tuberculosis | Blood | GSE89403 | RNA Grein | 23215 | Case = 100<br>Control = 38 | 271 | 182 | 89 |

### 3.2 Functional Pathway and GO Correlation Analysis

Five different databases were used for the pathway analysis: Elsevier Pathway, Bio Carta [31], Reactome [32], KEGG [33], and WikiPathway [34]. Then, we examined notably superior pathways utilizing DEGs among SARS-Cov-2, lung cancer, and tuberculosis. These pathways have been categorized into functional groups for ease of analysis. We determined 625 signaling pathways related to these diseases. Based on adjusted p-values <0.05, we selected manually the most significant pathways. We identified the most significant 55 transmission pathways. Using biological process (BP), molecular function (MF), and cellular component (CC) the GO procedure classified them into specific groups. Finally, we arranged the GO terms in ascending order by the adjusted p-value. We evaluated the databases of BP, MF and CC for GO terminologies. The most efficient GO keywords are adjusted p-value < 0.05. The most prominent ten listings in each of the biological techniques, cellular components, and molecular procedures are presented in Table 2. Table 3 depicts the top 20 pathways linked to TB, lung cancer, and SARS-Cov-2.

**Table 2:**
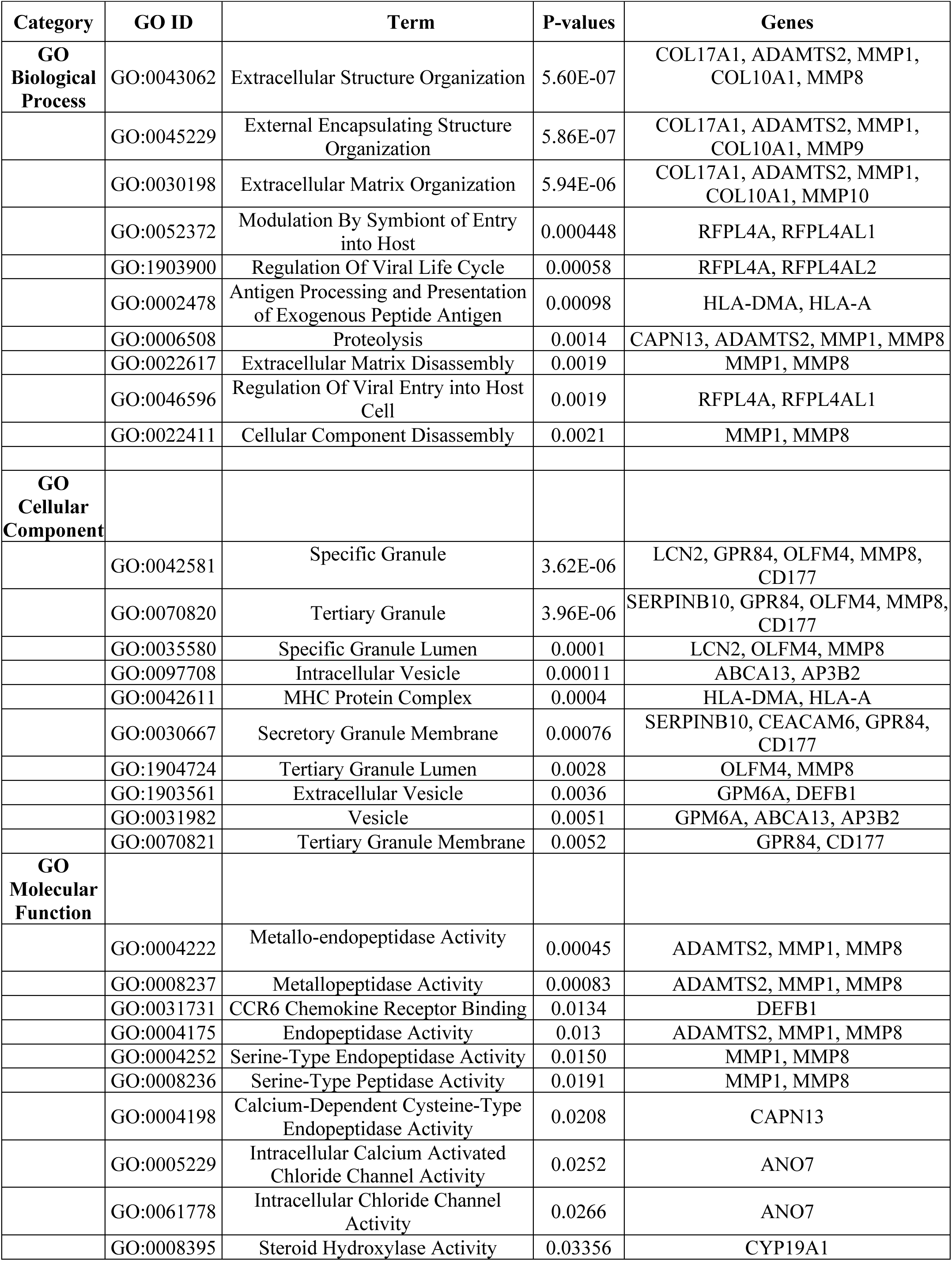
Ontological exploration of SARS COV-2, lung cancer, and tuberculosis DEGs.

| Category | GO ID | Term | P-values | Genes |
| --- | --- | --- | --- | --- |
| **GO Biological Process** | GO:0043062 | Extracellular Structure Organization | 5.60E-07 | COL17A1, ADAMTS2, MMP1, COL10A1, MMP8 |
|  | GO:0045229 | External Encapsulating Structure Organization | 5.86E-07 | COL17A1, ADAMTS2, MMP1, COL10A1, MMP9 |
|  | GO:0030198 | Extracellular Matrix Organization | 5.94E-06 | COL17A1, ADAMTS2, MMP1, COL10A1, MMP10 |
|  | GO:0052372 | Modulation By Symbiont of Entry into Host | 0.000448 | RFPL4A, RFPL4AL1 |
|  | GO:1903900 | Regulation Of Viral Life Cycle | 0.00058 | RFPL4A, RFPL4AL2 |
|  | GO:0002478 | Antigen Processing and Presentation of Exogenous Peptide Antigen | 0.00098 | HLA-DMA, HLA-A |
|  | GO:0006508 | Proteolysis | 0.0014 | CAPN13, ADAMTS2, MMP1, MMP8 |
|  | GO:0022617 | Extracellular Matrix Disassembly | 0.0019 | MMP1, MMP8 |
|  | GO:0046596 | Regulation Of Viral Entry into Host Cell | 0.0019 | RFPL4A, RFPL4AL1 |
|  | GO:0022411 | Cellular Component Disassembly | 0.0021 | MMP1, MMP8 |
| **GO Cellular Component** |  |  |  |  |
|  | GO:0042581 | Specific Granule | 3.62E-06 | LCN2, GPR84, OLFM4, MMP8, CD177 |
|  | GO:0070820 | Tertiary Granule | 3.96E-06 | SERPINB10, GPR84, OLFM4, MMP8, CD177 |
|  | GO:0035580 | Specific Granule Lumen | 0.0001 | LCN2, OLFM4, MMP8 |
|  | GO:0097708 | Intracellular Vesicle | 0.00011 | ABCA13, AP3B2 |
|  | GO:0042611 | MHC Protein Complex | 0.0004 | HLA-DMA, HLA-A |
|  | GO:0030667 | Secretory Granule Membrane | 0.00076 | SERPINB10, CEACAM6, GPR84, CD177 |
|  | GO:1904724 | Tertiary Granule Lumen | 0.0028 | OLFM4, MMP8 |
|  | GO:1903561 | Extracellular Vesicle | 0.0036 | GPM6A, DEFB1 |
|  | GO:0031982 | Vesicle | 0.0051 | GPM6A, ABCA13, AP3B2 |
|  | GO:0070821 | Tertiary Granule Membrane | 0.0052 | GPR84, CD177 |
| **GO Molecular Function** |  |  |  |  |
|  | GO:0004222 | Metallo-endopeptidase Activity | 0.00045 | ADAMTS2, MMP1, MMP8 |
|  | GO:0008237 | Metallopeptidase Activity | 0.00083 | ADAMTS2, MMP1, MMP8 |
|  | GO:0031731 | CCR6 Chemokine Receptor Binding | 0.0134 | DEFB1 |
|  | GO:0004175 | Endopeptidase Activity | 0.013 | ADAMTS2, MMP1, MMP8 |
|  | GO:0004252 | Serine-Type Endopeptidase Activity | 0.0150 | MMP1, MMP8 |
|  | GO:0008236 | Serine-Type Peptidase Activity | 0.0191 | MMP1, MMP8 |
|  | GO:0004198 | Calcium-Dependent Cysteine-Type Endopeptidase Activity | 0.0208 | CAPN13 |
|  | GO:0005229 | Intracellular Calcium Activated Chloride Channel Activity | 0.0252 | ANO7 |
|  | GO:0061778 | Intracellular Chloride Channel Activity | 0.0266 | ANO7 |
|  | GO:0008395 | Steroid Hydroxylase Activity | 0.03356 | CYP19A1 |

**Table 3:**
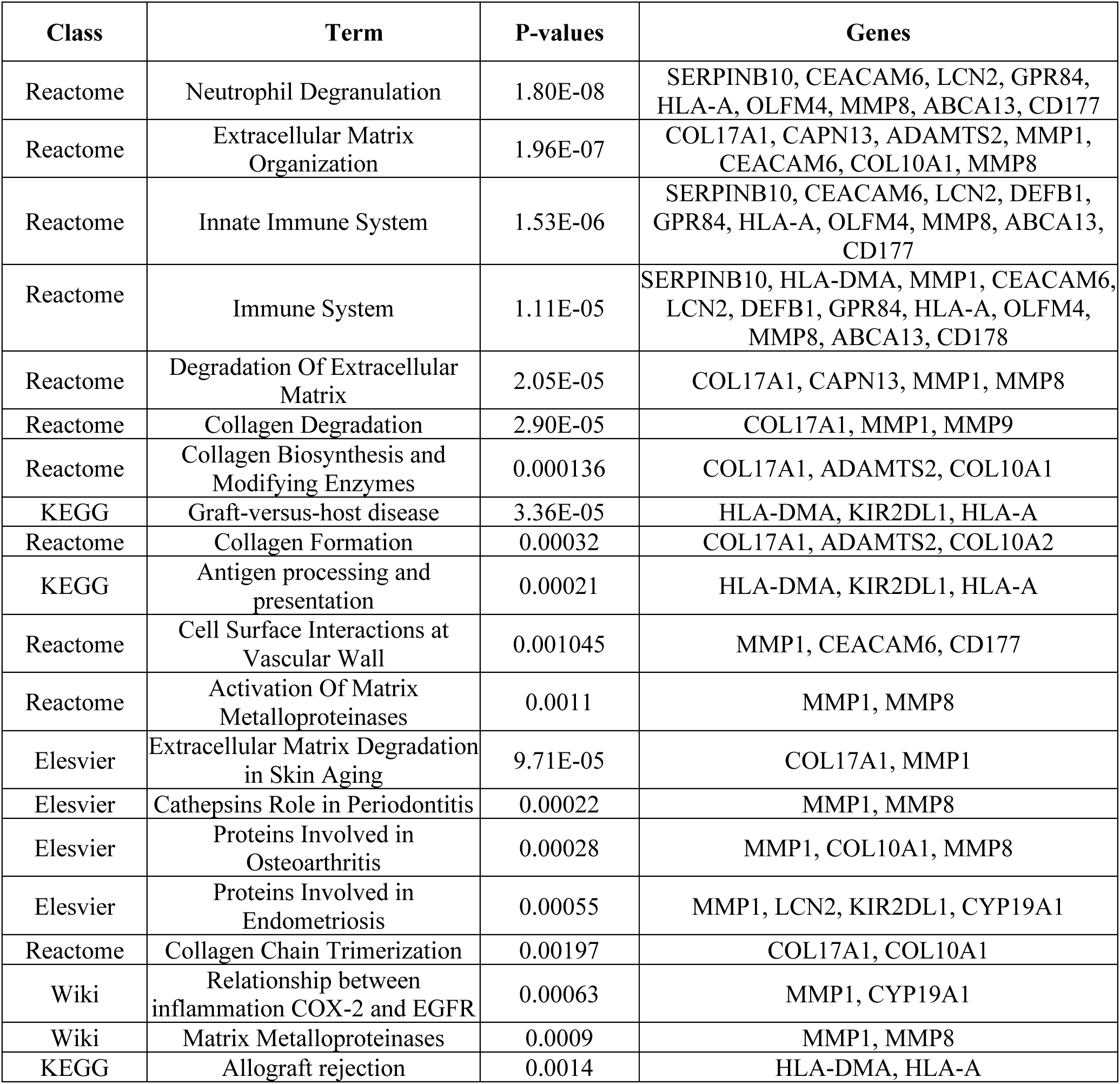
Pathways exploration of SARS COV-2, lung cancer, and tuberculosis DEGs.

| Class | Term | P-values | Genes |
| --- | --- | --- | --- |
| Reactome | Neutrophil Degranulation | 1.80E-08 | SERPINB10, CEACAM6, LCN2, GPR84, HLA-A, OLFM4, MMP8, ABCA13, CD177 |
| Reactome | Extracellular Matrix Organization | 1.96E-07 | COL17A1, CAPN13, ADAMTS2, MMP1, CEACAM6, COL10A1, MMP8 |
| Reactome | Innate Immune System | 1.53E-06 | SERPINB10, CEACAM6, LCN2, DEFB1, GPR84, HLA-A, OLFM4, MMP8, ABCA13, CD177 |
| Reactome | Immune System | 1.11E-05 | SERPINB10, HLA-DMA, MMP1, CEACAM6, LCN2, DEFB1, GPR84, HLA-A, OLFM4, MMP8, ABCA13, CD178 |
| Reactome | Degradation Of Extracellular Matrix | 2.05E-05 | COL17A1, CAPN13, MMP1, MMP8 |
| Reactome | Collagen Degradation | 2.90E-05 | COL17A1, MMP1, MMP9 |
| Reactome | Collagen Biosynthesis and Modifying Enzymes | 0.000136 | COL17A1, ADAMTS2, COL10A1 |
| KEGG | Graft-versus-host disease | 3.36E-05 | HLA-DMA, KIR2DL1, HLA-A |
| Reactome | Collagen Formation | 0.00032 | COL17A1, ADAMTS2, COL10A2 |
| KEGG | Antigen processing and presentation | 0.00021 | HLA-DMA, KIR2DL1, HLA-A |
| Reactome | Cell Surface Interactions at Vascular Wall | 0.001045 | MMP1, CEACAM6, CD177 |
| Reactome | Activation Of Matrix Metalloproteinases | 0.0011 | MMP1, MMP8 |
| Elesvier | Extracellular Matrix Degradation in Skin Aging | 9.71E-05 | COL17A1, MMP1 |
| Elesvier | Cathepsins Role in Periodontitis | 0.00022 | MMP1, MMP8 |
| Elesvier | Proteins Involved in Osteoarthritis | 0.00028 | MMP1, COL10A1, MMP8 |
| Elesvier | Proteins Involved in Endometriosis | 0.00055 | MMP1, LCN2, KIR2DL1, CYP19A1 |
| Reactome | Collagen Chain Trimerization | 0.00197 | COL17A1, COL10A1 |
| Wiki | Relationship between inflammation COX-2 and EGFR | 0.00063 | MMP1, CYP19A1 |
| Wiki | Matrix Metalloproteinases | 0.0009 | MMP1, MMP8 |
| KEGG | Allograft rejection | 0.0014 | HLA-DMA, HLA-A |

**Table 3:** Overview of SARS COV-2, lung cancer, and tuberculosis hub genes from protein-protein interactions.

| Gene Symbol | Description | Feature |
| --- | --- | --- |
| KIR3DL2 | Killer Cell Immunoglobulin Like Receptor, Three Ig Domains and Long Cytoplasmic Tail 2 | HLA-B27 dimers and free H chains have a higher affinity for binding compared to other HLA class I molecules. This binding facilitates the proliferation of T lymphocytes. |
| KIR3DL1 | Killer Cell Immunoglobulin Like Receptor, Three Ig Domains and Long Cytoplasmic Tail 1 | For HLA allele, receptor on natural killer (NK) cells. Suppresses the function of natural killer (NK) cells, hence inhibiting the destruction of cells. |
| HLA-DRA | Major Histocompatibility Complex, Class II, DR Alpha | *Peptide antigen binding and MHC class II receptor activity* |
| HLA-A | Major Histocompatibility Complex, Class I, A | *RNA binding and peptide antigen binding* |
| CD8A | CD8 Subunit Alpha | *Protein homodimerization activity and coreceptor activity* |
| LILRB1 | Leukocyte Immunoglobulin Like Receptor B1 | *Protein homodimerization activity and protein phosphatase 1 binding* |
| KIR2DL1 | Killer Cell Immunoglobulin Like Receptor, Two Ig Domains and Long Cytoplasmic Tail 1 | The receptor on natural killer (NK) cells is exclusive to certain HLA-C alleles. Restricts the function of natural killer (NK) cells, hence impeding the breakdown of cells. |
| KIR2DL3 | Killer Cell Immunoglobulin Like Receptor, Two Ig Domains and Long Cytoplasmic Tail 3 | Receptor on natural killer (NK) cells for HLA-C alleles. Inhibits the activity of NK cells thus preventing cell lysis |
| HLA-DRB1 | Major Histocompatibility Complex, Class II, DR Beta 1 | *Peptide antigen binding and MHC class II receptor activity* |
| HLA-C | Major Histocompatibility Complex, Class I, C | *Signaling receptor binding and TAP binding* |
| LCN2 | Lipocalin 2 | *Protein homodimerization activity and iron ion binding* |
| DEFB1 | Defensin Beta 1 |  |
| MMP8 | Matrix Metalloproteinase 8 | *Calcium ion binding and metallo-endopeptidase activity* |
| TIMP1 | TIMP Metalloproteinase Inhibitor 1 | *Cytokine activity and protease binding* |

### 3.3 Evaluation of hub proteins and module segments

Using the STRING database, We constructed a network of protein-protein interactions. The basis for this was differentially expressed genes that existed among SARS-CoV-2, TB and lung cancer. The results of this preparation and evaluation are displayed in Figure 4. The protein-protein interaction (PPI) network consists of a total of 26 nodes and 38 edges. Using the algorithms MCC and Degree, Figure 5 (A, B) displays the identified hub proteins. The algorithms were employed to identify a total of 22 hub genes. Among these, the top genes, which exhibited a high level of significance, were visually emphasized through the utilization of the 3 colors-red, orange, and yellow. Furthermore, using the MCC method, we identified 15 hub genes, of which 10 were the most significant (shown in Figure 5 as red, yellow, and orange colors). Hub genes are crucial components in a gene network due to their extensive interactions with other genes. Their importance extends to biological functions and gene regulation, and they are often strongly linked to diseases. Their ranking in the top 10% for connectivity further underlines their significance within the network. For example, if the module size was set at 100, the top-ranking ten genes would be selected as hub genes. The analysis of both methods yielded a selection of the top 14 Differentially Expressed Genes (DEGs) that exhibit the highest level of significance. These genes include KIR3DL2, KIR3DL1, HLA-DRA, HLA-A, CD8A, LILRB1, KIR2DL1, KIR2DL3, HLA-DRB1, HLA-C, LCN2, DEFB1, MMP8, and TIMP1. To understand the interconnections and proximity of hub genes, we constructed two submodule networks (Figure 5). The identification of these essential genes holds potential for their utilization as biomarkers, hence offering new avenues for therapeutic interventions in the studied diseases. The hub genes are outlined in Table 3.

**Figure 4:**
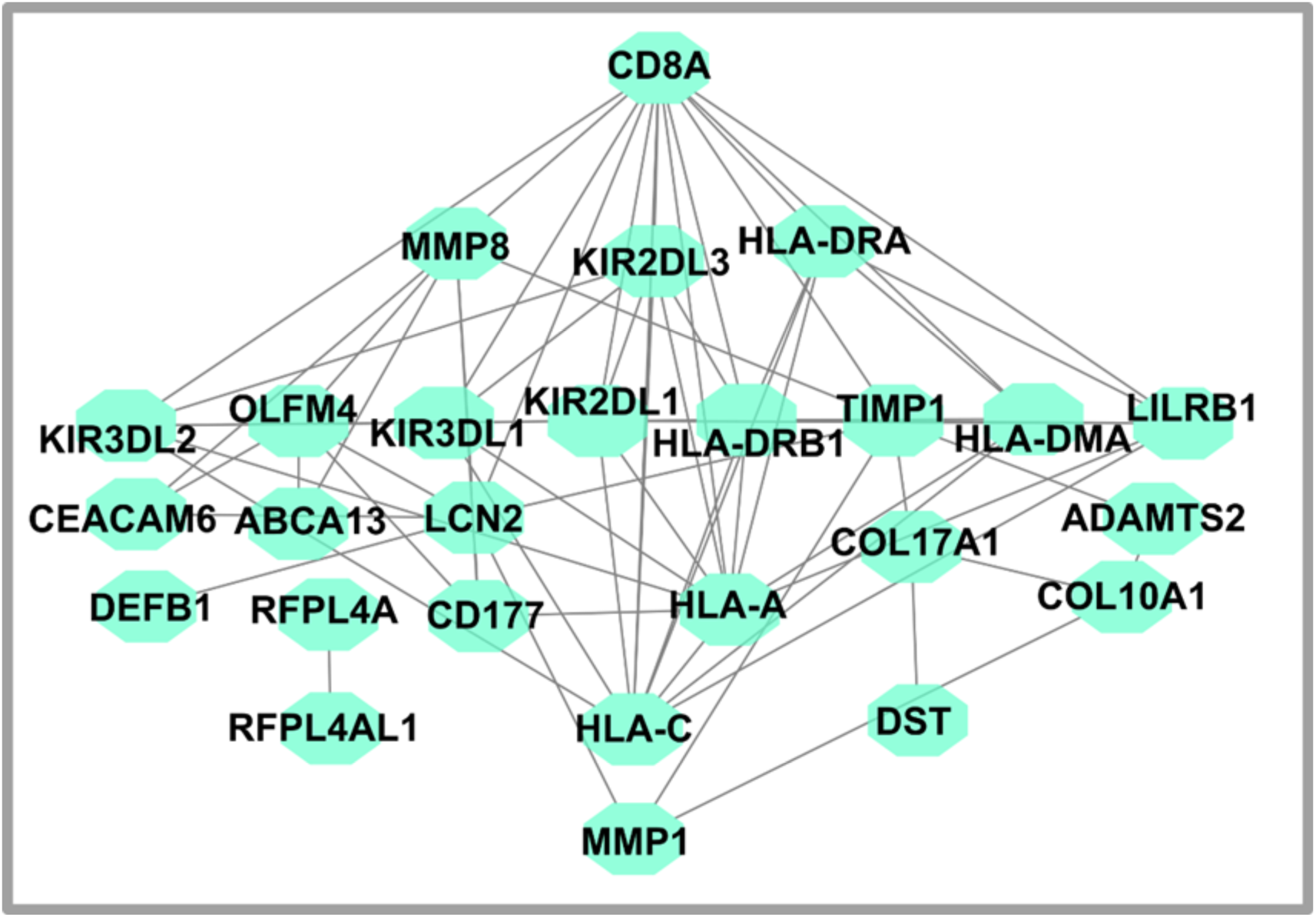
The network PPI of DEGs between SARS COV-2, tuberculosis and lung cancer, is illustrated in the figure. Denoted by circular nodes are DEGs, while edges represent node relations. With regard to the network of PPI, there are 26 nodes and 38 edges. The network of PPI was constructed and showed in Cytoscape using the String library.

**Figure 5:**
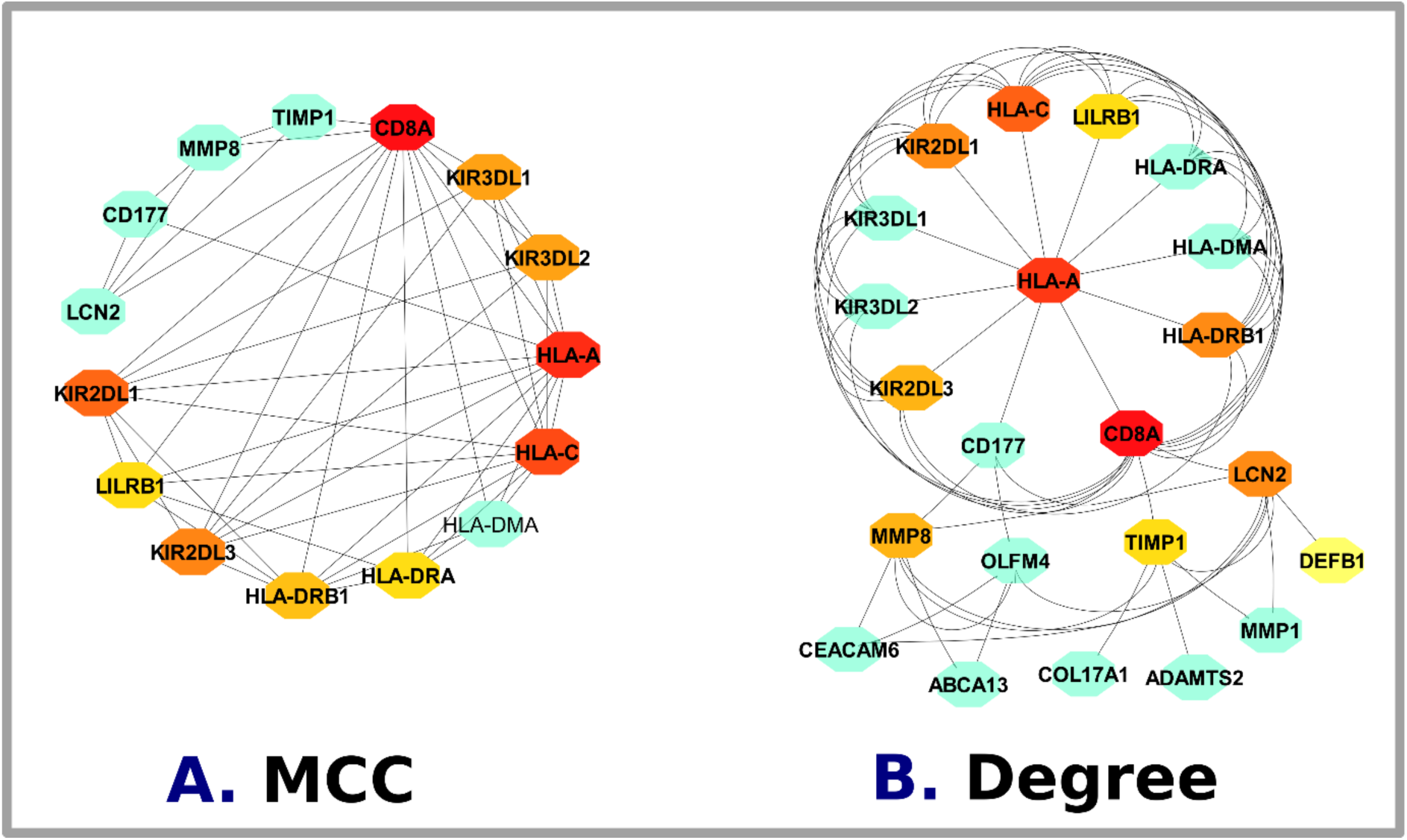
Identifying the network of PPI hub genes with Cytohubba in Cytosacpe. The latest MCC and Degree Cytohubba plugins were used to find hub genes.

### 3.4 Identification of the Differentially Expressed Genes’ Transcriptional and Post-Transcriptional Regulators

The control of gene expression is greatly impacted by transcription factors, and illnesses can emerge when these factors are dysregulated or exhibit aberrant activity. Proteins called transcription factors help convert DNA to RNA, a process known as transcription [52]. Transcription is the process by which genes are transcribed into RNA or proteins [53]. Excluding RNA polymerase, transcription factors comprise a large number of proteins that initiate and regulate the transcription of genes [54]. Transcription factors, which are found in all living creatures, influence gene expression [55]. Many biological processes are controlled by TF genes, which makes them very important [56]. MicroRNAs are highly conserved molecules of noncoding RNA that regulate gene expression [57]. MicroRNA mostly controls gene expression by forming a bond with messenger RNA (mRNA) in the plasma membrane of the cell [58][59]. Insufficient amounts of a microRNA may lead to the over activation of genes that it controls, hence facilitating the growth and advancement of cancer [60]. It has been discovered that miRNAs are dysregulated throughout the start and growth of lung cancer and may regulate lung cancer cell proliferation and invasion [61]. The pathogenesis of SARS CoV-2 has been associated with miRNA dysregulation in numerous studies, including viral replication and entrance, inflammatory response, ACE2, and cytokine storm, and miRNA dysregulation to the pathogenesis of tuberculosis, including immune modulation and granuloma formation [62]. Through an investigation of the interaction between TFs and DEGs, we explored the post-transcriptional and transcriptional regulators of DEGs and the results shown in Fig. 6 and the miRNAs-DEGs reaction study shown in Figure 7. Figure 6 shows the relationship between TFs and common genes of SARS-CoV-2, tuberculosis, and lung cancer. The regulating genes are: GATA2, NFKB1, FOXL1, STAT3, FOXC1, SRF, YY1, USF2, GLIS2, PHF2, DPF2, SIN3A, ZFP37, FOXM1, ZNF580, SP2, CTCF, SMAD4, EZH2, ZNF263, KLF4. Figure 8 shows that the mRNA related with common genes of SARS CoV-2, tuberculosis, and lung cancer: mir-124-3p, mir-335-5p, mir-767-5p, mir-29a-3p, mir-219-5p, mir-27a-3p, mir-148b-3p, mir-1-3p, mir-27a-5p, mir-26a-5p, mir-34a-5p, mir-200b-3p, mir-1343-3pA summary of post-transcriptional regulating biomolecules associated with Type 2 Diabetes (T2D) and breast cancer is shown in the following Table 4.

**Figure 6:**
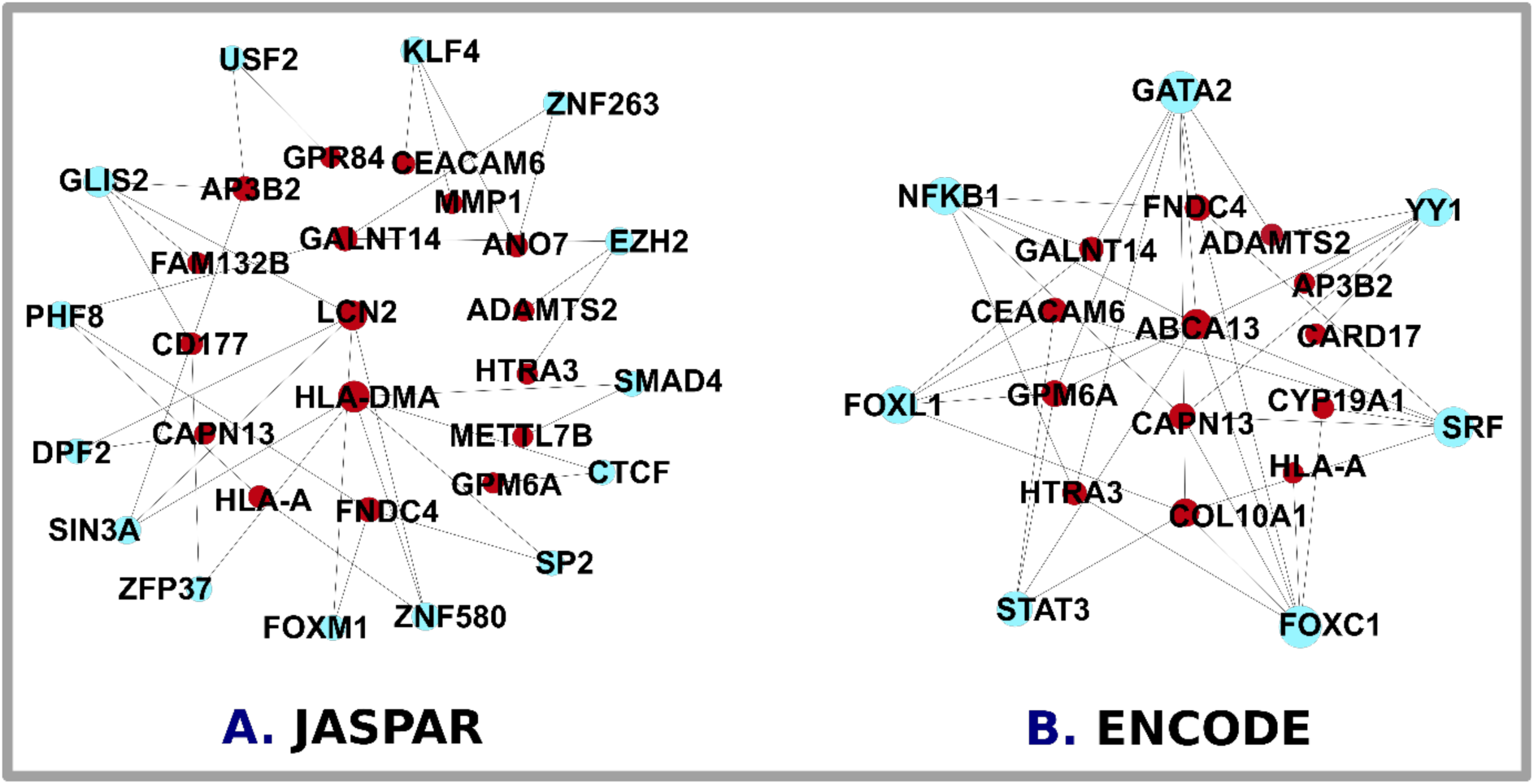
TF-Gene correlations from JASPAR and ENCODE showed the link between SARS COV-2, lung cancer, and tuberculosis. The blue circles represent DEGs gene-interacting TF-genes.

**Figure 7:**
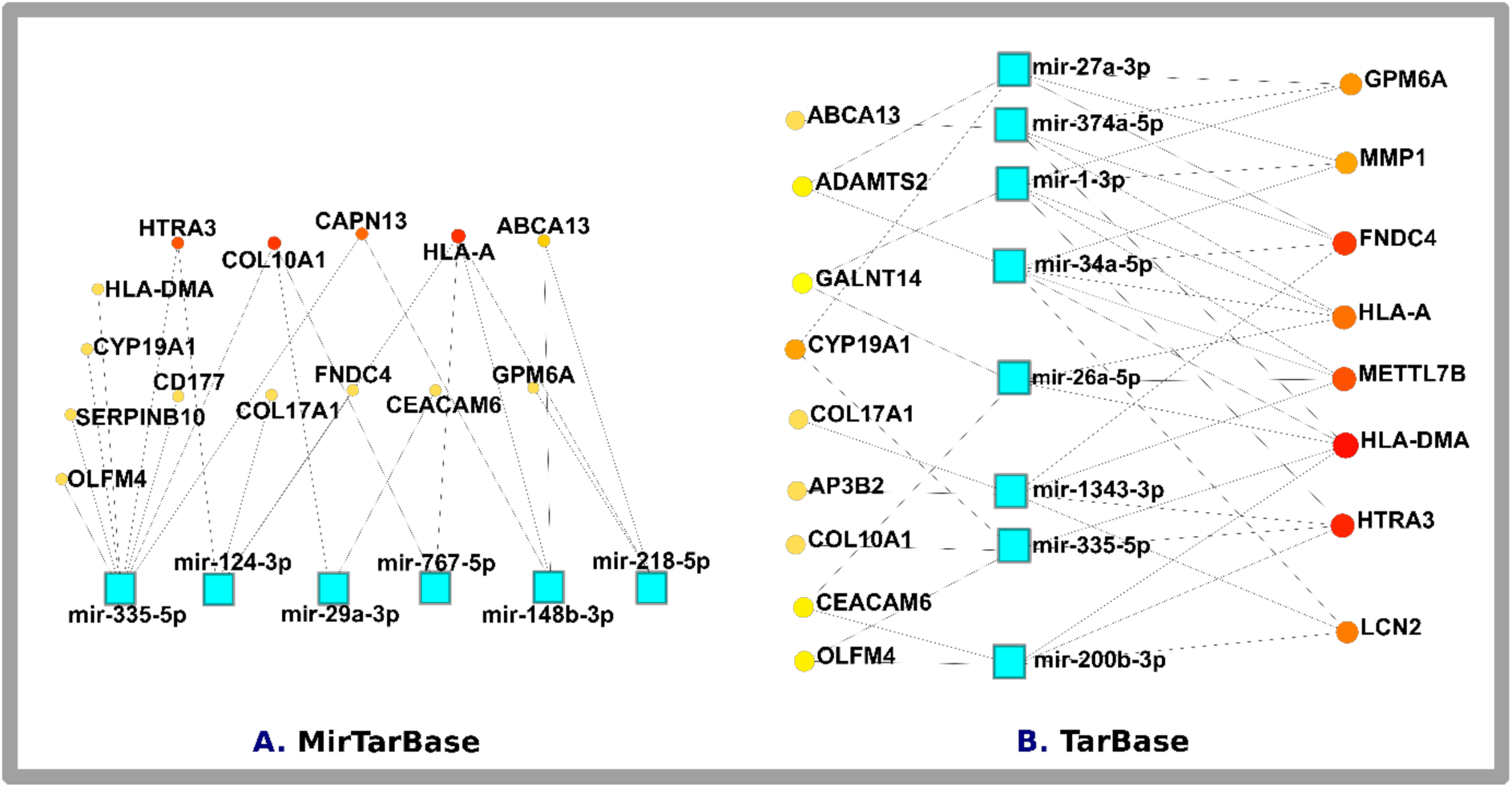
The miRNA gene in SARS COV-2, lung cancer, and T2D was found using TarBase and MirTarBase. The circular yellow colors, orange, and red (low moderate and high) indicate DEGs genes interacting with the mi-RNA gene (blue square box).

**Figure 8:**
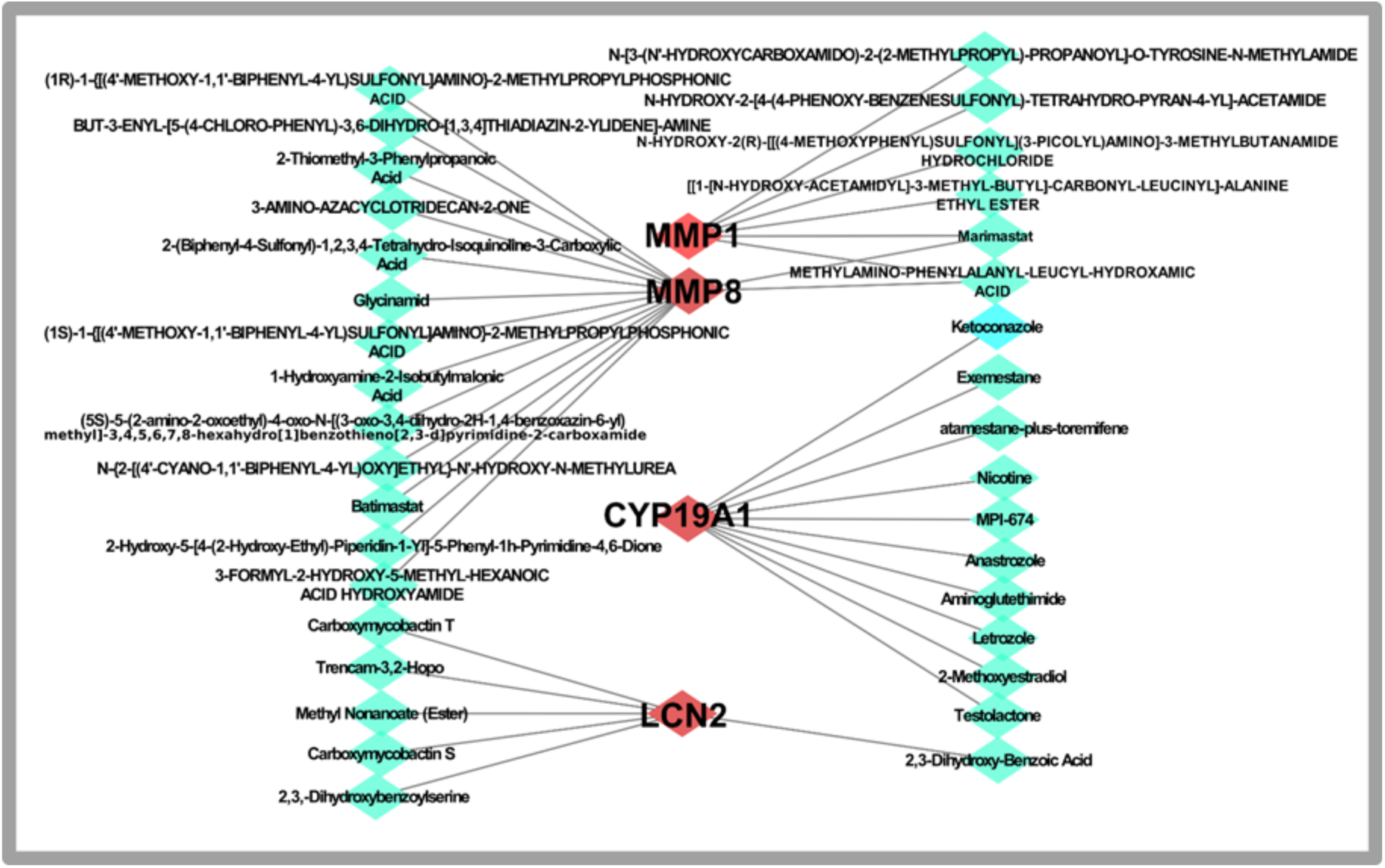
The figure illustrates the Protein Drug Interaction technique, showcasing the interaction between proteins and drugs.

**Table 4:** Summary of SARS-CoV-2, lung cancer, and tuberculosis post-transcriptional regulatory biomolecules.

| Symbol | Description | Feature |
| --- | --- | --- |
| mir-335-5p | MicroRNA 335 | Controls the growth, movement, and infiltration of cancer cells. |
| mir-124-3p | MicroRNA 124 | Suppresses the advancement of lung cancer by targeting the PI3K/AKT signaling cascade |
| mir-29a-3p | MicroRNA 29a | Controlled several fundamental characteristics of cancer, including as cellular proliferation, movement, and the development of tumors |
| mir-767-5p | MicroRNA 767 | Blocks the growth and spread of glioma by specifically targeting SUZ12 |
| mir-148b-3p | MicroRNA 148b | Suppresses the transmission of gastric cancer by blocking the Dock6/Rac1/Cdc42 pathway |
| mir-219-5p | MicroRNA 219a-2 | Inhibits the growth and advancement of cells in esophageal squamous cell carcinoma by specifically targeting CCNA2. |
| mir-27a-3p | MicroRNA 27a | Changing the malignant, biological action of pancreatic cancer cells by targeting Spry2 |
| mir-27a-5p |  |  |
| mir-1-3p | MicroRNA 1 | The process of mesenchymal differentiation of stem cells is controlled in order to avoid the development of osteoporosis. This is achieved by specifically targeting a protein called secreted frizzled-related protein. 1 Controlling cancer-related mechanisms, such as the growth of cells, programmed cell death, the transition of cells from epithelial to mesenchymal state, and the spread of cancer to other parts of the body |
| mir-34a-5p | MicroRNA 34a | Controlling cancer-related mechanisms, such as the growth of cells, programmed cell death, the transition of cells from epithelial to mesenchymal state, and the spread of cancer to other parts of the body |
| mir-26a-5p | MicroRNA 26a | Inhibits apoptosis and inflammatory responses via targeting CTGF in A549 cells |
| mir-1343-3p | MicroRNA 1343 | Inhibit the activation of TGF-β receptor genes and prevent the transformation of epithelial cells into mesenchymal cells caused by TGF-β. |
| mir-200b-3p | MicroRNA 200b | The Wnt/$\beta$ catenin signaling pathway was deactivated, resulting in suppressed cell proliferation and increased cell death by apoptosis in colorectal cancer (CRC) cells. |

**Table 4:** Summary of SARS-CoV-2, lung cancer, and tuberculosis transcriptional regulatory biomolecules.

| Gene Symbol | Description | Feature |
| --- | --- | --- |
| GATA2 | GATA Binding Protein 2 | *chromatin binding and DNA-binding transcription factor activity chromatin binding* |
| NFKB1 | Nuclear Factor Kappa B Subunit 1 | *sequence-specific DNA binding and DNA-binding transcription factor activity and* |
| FOXL1 | Forkhead Box L1 | *DNA-binding transcription factor activity, RNA polymerase II-specific and DNA-binding transcription factor activity and* |
| STAT3 | Signal Transducer and Activator of Transcription 3 | *sequence-specific DNA binding and DNA-binding transcription factor activity* |
| FOXC1 | Forkhead Box C1 | *transcription factor binding DNA-binding and transcription factor activity* |
| SRF | Serum Response Factor | *DNA-binding transcription factor activity and sequence-specific DNA binding* |
| YY1 | YY1 Transcription Factor | *transcription coactivator activity and DNA-binding transcription factor activity* |
| USF2 | Upstream Transcription Factor 2, C-Fos Interacting | *sequence-specific DNA binding and DNA-binding transcription factor activity* |
| GLIS2 | GLIS Family Zinc Finger 2 | *transcription cis-regulatory region binding and DNA-binding transcription activator activity, RNA polymerase II-specific* |
| PHF2 | PHD Finger Protein 2 | *transcription coactivator activity and methylated histone binding* |
| DPF2 | Double PHD Fingers 2 | *RNA polymerase II cis-regulatory region sequence-specific DNA binding* |
| SIN3A | SIN3 Transcription Regulator Family Member A | *transcription factor binding and DNA-binding transcription factor activity* |
| ZFP37 | ZFP37 Zinc Finger Protein | *DNA-binding transcription factor activity and Nucleic acid binding* |
| FOXM1 | Forkhead Box M1 | *protein kinase binding and DNA-binding transcription factor activity* |
| ZNF580 | Zinc Finger Protein 580 |  |
| SP2 | Sp2 Transcription Factor | *histone deacetylase binding and DNA-binding transcription factor activity, RNA polymerase II-specific* |
| CTCF | CCCTC-Binding Factor | *chromatin binding and DNA-binding transcription factor activity* |
| SMAD4 | SMAD Family Member 4 | *sequence-specific DNA binding and DNA-binding transcription factor activity* |
| EZH2 | Enhancer Of Zeste 2 Polycomb Repressive Complex 2 Subunit | *chromatin binding and Sequence-specific DNA binding* |
| ZNF263 | Zinc Finger Protein 263 | *DNA-binding transcription factor activity and Nucleic acid binding* |
| KLF4 | KLF Transcription Factor 4 | *transcription factor binding and DNA-binding transcription factor activity* |

### 3.5 Identification of potential drugs

Assessing the relationship between medications and proteins is a vital process for comprehending the effectiveness and consequences of the drug [63]. Drug discovery requires the identification of novel drug–protein interactions. Protein-drug connections are vital to comprehending ligand affinity [64] [65]. For SARS-CoV-2, lung cancer, and tuberculosis, we determined 35 potential therapeutic compounds for prevalent DEGs as potential drugs. Figure 8 depicts Protein Drug Interactions for SARS-CoV-2, Lung cancer, and Tuberculosis complications based on the MMP1, MMP8, CYP19A1, and LCN2 genes. The connections between MMP1, MMP8, CYP19A1, and LCN2 and drug targets were the strongest, so we focused on this gene as a potential therapeutic candidate.

## 4. Discussion

The SARS-CoV-2 (COVID-19) virus mostly affects the respiratory system and, in certain cases, can result in severe pneumonia [66]. It can worsen pre-existing respiratory illnesses, such as lung cancer and tuberculosis, thereby increasing the susceptibility of people with these conditions to poor consequences [67]. SARS-CoV-2, the virus causing COVID-19, has the capacity to impair the immune system in certain individuals, particularly those with severe manifestations of the disease [68]. Immunosuppression increases their vulnerability to other illnesses, such as tuberculosis (TB) [69]. Uncontrolled proliferation of cells within the pulmonary system is the hallmark of lung cancer [70]. The condition can manifest as a chronic cough and difficulty breathing, which may be mistaken for symptoms of COVID-19, resulting in delays in identifying and treating the underlying issue [71]. Lung cancer, along with other types of cancer, has the potential to compromise the immune system [72].

Cancer therapies such as chemotherapy and radiation can also inhibit the immune system, rendering patients more susceptible to infections, especially respiratory infections such as COVID-19 and TB [73]. Lungs are the main organs affected by tuberculosis (TB), an infectious disease [74]. It has the potential to compromise the immune system and increase vulnerability to other respiratory diseases, such as COVID-19 [75]. Tuberculosis itself compromises the immune system, rendering individuals more vulnerable to other illnesses, such as SARS-CoV-2 [76]. Symptoms that are shared by COVID-19, lung cancer, and tuberculosis include cough, fever, and fatigue [77]. This might result in diagnostic complexities, as healthcare personnel may be required to distinguish between these illnesses [78]. The COVID-19 pandemic’s diversion of healthcare resources to address the epidemic’s excessive number of COVID-19 cases may have impacted the diagnosis and treatment of TB and lung cancer patients [79].

Gene ontology (GO) enhancement analysis generates ideas about the fundamental physiological characteristics of research and assesses large-scale molecular data [80]. We initially focused on the main GO biological procedure [81]. The ECM, or extracellular matrix, is a complecated network made of many extracellular macromolecules and nutrients, such as elastin and collagen, proteoglycans, glycosaminoglycans, laminins, fibronectin, glycoproteins, enzymes, and hydroxyapatite, that serves both structural and biochemical protection to the adjacent cells [82]. COVID-19 exhibits a systemic inflammatory reaction and atypical expression of the extracellular matrix (ECM), which governs homeostasis and response to injury [83]. The extracellular matrix is a constantly changing part of the tumor microenvironment that controls the ability of tumor cells to move away from the original tumor and spread to nearby or distant locations [84]. Symbiosis refers to a close connection between individuals of distinct species, such as the interactions between complex multicellular animals and microorganisms. The microorganisms that form mutually beneficial relationships with other organisms are referred to as symbionts [85]. Symbiotic helminths and protists were connected to a decreased likelihood of experiencing severe symptoms of Coronavirus Disease 2019 (COVID-19) [86]. The microbiome significantly impacts human health and is implicated in the development and advancement of lung cancer by triggering inflammatory responses, influencing immunological control, and contributing to metabolic abnormalities and genotoxicity [87]. Tuberculosis is a microorganism that may behave as both a disease-causing agent and a symbiont, which has significant implications for our comprehension of how hosts and pathogens interact [88]. The process of virus penetration into animal tissues begins with the attachment of the virus to receptors, that is followed by significant structural modifications in viral proteins, allowing The virus may either permeate the membranes of cells (in instances of non-enveloped viruses) or combine with cellular membrane (enveloped viruses) [89]. Coronaviruses gain entry into host cells by utilizing a trimeric spike protein that is connected to the envelope. This spike protein attaches to a receptor on the host cell, initiating the fusion of the viral and host cell membranes. The spike protein comprises of receptor-binding (S1) and membrane-fusion (S2) domains [90]. The human papilloma virus (HPV), which is known to cause genital malignancies, has also been proposed as a potential factor in the development of lung cancer [91]. Mycobacterium tuberculosis (Mtb) is an exceptionally thriving pathogen, specifically suited for humans and responsible for tuberculosis. The effectiveness of the pathogen is linked to its capacity to suppress the innate immune responses of the host cell through the utilization of a variety of virulence factors with distinct characteristics [92]. Proteolysis refers to the process by which a protein is broken down, either partly into peptides or totally into amino acids. This breakdown is facilitated by proteolytic enzymes, which are found within bacteria, plants, and particularly in animals [93]. The primary coronavirus protease, known as SARS-CoV-2 Mpro or 3CLpro, has a crucial function in the cleavage of polyproteins synthesised from viral RNA during the replication phase of SARS-CoV-2 [94]. More precisely, researchers have developed tiny-molecule proteolysis-attacking chimaeras (PROTACs) to target specific proteins in cancer cells. These PROTACs consist of two ligands joined by a linker, which binds to the target protein, and an E3 ubiquitin ligase. This development has shown crucial possibilities for treating progressive lung cancer [95]. The Mycobacterium TB genome contains 142 proteases, including FtsH, Clp proteases, and the proteasome, which are responsible for causing tuberculosis [96]. Additionally, our attention was directed towards cellular components, namely the prominent Gene Ontology (GO) terms: Specific Granule, Tertiary Granule, Extracellular Vesicle, Intracellular Vesicle, and MHC Protein Complex.

Intracellular granules are little particles that may be challenging to observe using a conventional light microscope. The term “granule” is commonly used to refer to secretory vesicles located solely in immune cells known as granulocytes [7]. Granulocytes, known as granular leukocytes, are the predominant kind of white blood cells. Granulocytes, also known as granular leukocytes, can be further classified based on the staining qualities of their granules. Neutrophils, eosinophils, and basophils Neutrophils possess a minimum of four distinct types of granules: (1) primary granules, also referred to as azurophilic granules; (2) secondary granules, also termed particular granules; (3) tertiary granules; and (4) secretory vesicles [98]. Following infection with SARS-CoV-2, an increased quantity of neutrophils has been detected in the nasopharyngeal epithelium and subsequently in the deeper regions of the lungs. Severe cases of COVID-19 exhibit notable upregulation of markers associated with immature or early-stage neutrophils, in contrast to mild cases [99]. Neutrophils and neutrophil extracellular traps (NETs) have a role in this elevated risk, and many subgroups of neutrophils have been identified in lung cancer patients [100]. Granulocytes have the ability to behave as MDSCs(myeloid-derived suppressor cells) that can hinder a defensive adaptive immune response by suppressing the activity of T-cells in the context of Mycobacterium TB infection [101]. A vesicle is a membrane structure that stores and transfers biological products and breaks down metabolic waste in the cell. Vesicles are formed by the cell during exocytosis, endocytosis, and intracellular trafficking [102]. During the COVID-19 infection, extracellular vesicle (EVs) has a role in promoting blood clot formation, which leads to impaired blood vessel function. This process exacerbates the excessive release of cytokines, known as hypercytokinemia, resulting in a severe immune response called a cytokine storm [103]. Enhancing our comprehension of extracellular vesicle (EV)-mediated intercellular communication and the contents of EVs could be highly beneficial for the spread of lung cancer to other parts of the body and identifying clinical indicators [104]. Mycobacterium tuberculosis (M. tuberculosis) generates extracellular vesicles(EVs) both in laboratory conditions and within living organisms. These EVs contain membrane-bound nanoparticles that aid the transfer of various biological substances such as lipids, nucleic acids, proteins, and glycolipids. Additionally, these EVs can interact with the host organism from a distance [105]. The Major Histocompatibility Complex (MHC) type I and type II proteins have a crucial function in the adaptive part of the immune system [106]. The interaction between MHC I epitope content and the SARS-CoV-2 peptide directly impact the ability of T cells to defend against infection. Thus, the process of MHC presentation is strongly linked to the condition of lymphopenia in individuals with COVID-19 [107]. Throughout the process of cancer development and progression, MHC-I molecules display tumor-associated antigens, which are identified as foreign pathogens. This recognition stimulates the destruction of tumor cells by cytotoxic T lymphocytes [108]. Mycobacterium tuberculosis thrives within antigen-presenting cells (APCs), such as macrophages and dendritic cells. Antigen-presenting cells (APCs) display antigens alongside major histocompatibility complex (MHC) type II molecules to activate CD4+ T cells, which is crucial for controlling M. tuberculosis infection [109]. An endopeptidase is an enzyme that cleaves peptide bonds within a protein molecule. Proteins undergo peptide chain fragmentation as a consequence of the endopeptidase process [110]. The SARS-CoV-2 virus binds to angiotensin-converting enzyme 2 (ACE2) receptors on the membrane of the host cell, facilitated by dipeptidyl peptidase 4 (DPP4), which are both types of peptidases, with DPP4 being an exopeptidase and ACE2 being an endopeptidase [111]. This endopeptidase enzymatically breaks down bioactive peptides, such as bombesin-like peptides, as well as related neuropeptides, through hydrolysis. Bombesin-like peptides and other neuropeptides function as autocrine growth agents for both small-cell lung cancer (SCLC) and non-small-cell lung cancer (NSCLC) [112]. The process of cell separation relies on the activity of cell-wall hydrolases, which break down the peptidoglycan layer that links the daughter cells together. The regulation of this mechanism in Mycobacterium tuberculosis is controlled by the anticipated endopeptidase RipA. Without this enzyme, the bacteria cannot undergo cell division and display an unusual phenotype [113]. Chemokines, also known as chemotactic cytokines, are a diverse group of tiny proteins that are produced and transmit signals through cell surface G protein-coupled hepta-helical cytokine receptors. Chemokines have a crucial function in the formation and maintenance of the immune system and participate in all immunological and inflammatory reactions that can either protect or harm the body [114]. Chemokines may serve as the primary catalyst for acute respiratory distress syndrome, a significant consequence that resulted in mortality in almost 40% of severe cases during the recent COVID-19 pandemic. Multiple clinical studies have demonstrated that chemokines play a direct role in various phases of SARS-CoV-2 infection [115]. Chemokines and their receptors, which are essential components in the microenvironment, have significant impacts on the development, advancement, and spread of lung tumors. Investigating the processes behind this phenomenon could provide valuable knowledge for early detection and targeted treatment [116]. Chemokines have a crucial function in coordinating the recruitment of cells into the lung infected with Mtb, which aids in containing Mycobacterium tuberculosis [117]. Chloride channels play a crucial role in various biological processes such as epithelial fluid release, cell volume control, neuroexcitation, smooth muscle contraction, and the acidification of organelles within cells [118]. Studies have shown that ion channel activity has a role in controlling cell death (apoptosis) in several diseases. Therefore, identifying methods to regulate ion channel activity could provide a novel approach to preventing apoptosis mediated by SARS-CoV [119]. Ion channels, including Ca2+-activated chloride channels, play a significant role in controlling cell proliferation, cell movement, and metastasis. They are considered crucial targets for developing drugs to treat lung cancer [120].

A sequence of genes in the module pointed to as the most closely connected and possessing greater biological impact is most strongly associated with the hub genes [121]. (KIR) are a diverse group of receptors that play a crucial role in regulating the function of human NK cells. These receptors can either activate or inhibit the function of NK cells and exhibit a high degree of genetic variation. The functional role of various KIR family members is determined by the presence of unique structural domains, which serve as specific binding sites for ligands or signal proteins [122]. Several viral infections have cleared up by inheriting both the cognate ligand HLA-Bw4 and the inhibitory KIR3DL1 gene. The KIR3DL1+HLA-Bw4+ combination provides protection against several viral diseases, such as SARS-CoV2, HIV, H1N1/09, and human papillomavirus [123]. The unique combination of killer cell immunoglobulin-like receptors and corresponding HLA class I ligands that regulates natural killer cell immunity to viral infections makes individuals more susceptible to certain diseases, COVID-19. Frontiers in Genetics, 13, 845474. KIR genes and HLA-I ligands that specify the educational level of natural killer (NK) cells may influence the risk of developing lung cancer, according to our findings [124]. Cancer cells and TILs exhibited expression of KIR2D (L1, L3, L4, S4) and KIR3DL1, according to a study involving 62 patients with lung cancer. The high expression of KIR2D on cancer cells correlated with the high expression of KIR3DL1 and KIR2D on TILs [125]. Activating genes, KIR2DS5 KIR2DS1, and inhibitory genes KIR2DL3and KIR3DL1 bestowed susceptibility to tuberculosis either singly or in combination with other haplotypes [126]. The modulation of disease severity in HIV-1 and tuberculosis infections is facilitated by the expression of KIR3DS1/L1, which is associated with IFN-/IL-10 responses [127]. The viral antigen presentation pathway is significantly influenced by human leukocyte antigen (HLA), which is a critical element in determining disease severity and viral susceptibility. The immune response has been influenced by HLA alleles in viral diseases, including SARS-CoV-2 [128]. Presented as cellular antigens to T cells, classic type I HLA molecules (HLA-A, HLA-B, and HLA-C) are indispensable for cancer immunotherapy and immune surveillance [129].

Deregulation of HLA-I in cancer and its critical role in immunotherapy Journal of Cancer Immunotherapy, 9(8) Lung cancer frequently exhibits a dysregulation or absence of human leukocyte antigens (HLAs), which is a frequent indicator of carcinogenesis since cancer immune surveillance is predominately dependent on immunogenic antigens [130]. Human leukocyte antigen (HLA) class II alleles are essential for the early immune response to tuberculosis (TB) by presenting antigenic peptides to CD4+ T cells. Consequently, variations in these genes can impact the efficacy of the immune response to infection and the advancement of active disease [131]. CD8 T lymphocytes constitute a critical component of the antiviral immune system. Effector CD8 T lymphocytes make antiviral cytokines like interferon-gamma and eliminate virus-infected cells. CD8 T lymphocytes thereby aid in the resistance against both primary and secondary infection with viruses [132]. A SARS-CoV-2-induced illness or malignancy is indicated by dysregulated innate and adaptive immunity.

CD8+ T cells are essential immune system cells. As components of adaptive immunity, these cells serve as the primary line of protection against malignancy and viral infections. Severe CD8+ T-cell activation in the lung and tumor microenvironment (TME) of patients with SARS-CoV-2-induced disease will cause these cells to enter an exhausted state and endure apoptosis [133]. The function performed by CD8+ T cells in opposition to tumor cells is vital. Nevertheless, as stated previously, lung tumor cells elicit a sequence of qualitative and quantitative modifications in CD8+ T-cells, which impede their complete engagement in the identification and eradication of the tumor [134]. The vital parts of the host’s TB defense are CD4+ and CD8+ T cells [135]. CD8+ T cells are crucial for effective control of Mycobacterium tuberculosis, as they directly eliminate infected host cells and promote long-lasting immunological memory, according to accumulating evidence. Humans fail to generate a robust CD8+ T cell memory in response to Mycobacterium tuberculosis infection; similar results have been observed in animal models, even after successful treatment [136]. It was observed that the capacity of natural killer (NK) cells derived from COVID-19-positive patients to lyse SARS-Cov-2-infected cells is significantly enhanced when blocking antibodies specific for receptors KIR2DL1 and NKG2A are utilized [137]. The characterization of NK cells that infiltrate NSCLC for the first time reveals that their primary function is the production of pertinent cytokines, not direct cancer cell destruction. In order to determine whether or not NK cells infiltrating human non-small cell lung cancers (NSCLC) play a protective function in an antitumor immune response, and their activity was analyzed. Comparative analyses were conducted on the functions and expression of pertinent molecules of NK cells that infiltrate NSCLC and Autologous natural killer (NK) cells obtained from normal lung tissues around the tumor or from peripheral blood [138]. An upregulation of the KIR A haplotype was observed in tuberculosis patients relative to the control group.

Among TB patients, only KIR 2DL3 exhibited a substantially higher prevalence [139]. Infection-related immune system responses and the interaction between the host and pathogen are significantly influenced by human leukocyte antigens (HLA) [140]. As the host HLA is central to the activation and regulation of the immune response, the clinical trajectory of SARS-CoV-2 infection is highly dependent on the virus’s interaction with the host immune system [141]. HLA-G, which is expressed by the most prevalent malignant tumors worldwide (colorectal, gastric, esophageal, and lung cancer), is a tumor-associated molecule.

Subsequently, it is targeted by GC, ESCC, NSCLC, and colorectal cancer [142]. In an attempt to establish a correlation between pulmonary tuberculosis and HLA specificities, numerous researchers have undertaken population studies. However, no discernible association has been found between pulmonary TB and any particular HLA-A, -B, or specificity or HLA haplotype [143].

In addition, we analyzed the connections between TF genes and miRNAs to determine the regulatory factors involved in the transcriptional and post-transcriptional regulation of the common DEGs. Transcription factors (TFs) regulate the transcription ratio, while microRNAs (miRNAs) are vital in regulating genes and silencing RNA at the post-transcriptional level. TFs and miRNAs are important to understand disease initiation. Thus, our investigation established a connection between the common DEGs, TFs, and miRNAs. We identified multiple TF genes, including FOXC1, GATA2, YY1, SIN3A, and NFKB1. FOXM1 plays a crucial role in the progress of the cell cycle & overexpression of FOXM1 can prevent lung cancer cell apoptosis and enhance their proliferation by controlling the transitions between the G1/S and G2/M phases and maintaining the accurate implementation of mitotic cell division [144] [145]. GATA2 is a member of the GATA family of zinc finger proteins. It functions as a transcriptional regulator, either activating or repressing many genes that control a wide range of cellular activities in both development and carcinogenesis [146]. GATA2 has been classified as a gene susceptible to epigenetic silencing in cells of non-small cell lung cancer (NSCLC) [147]. Downregulation of GATA2 inhibited the improvement of TMPRSS2 transcription in response to IL1β interaction. The IL1β-promote TMPRSS2 overexpression could increase the cellular risk of SARS-CoV-2 infection [148]. In the lungs of experimental TB cases, YY1 is overexpressed. Treatments that inhibit YY1 expression could represent an innovative therapeutic approach to tuberculosis [149]. However, in lung cancer, the downregulation of the YY1 transcriptional axis promotes neo-angiogenesis and cancer progression [150]. Overexpression of SIN3A levels in plasma and skin microvasculature was linked to COVID-19 severity, Dysregulation of SIN3A may lead to the disruption of normal gene regulation in growth-related genes via histone acetylation, which in turn stimulates the development of lung cancer cells [151][152]. NF-κB1 is a member of NF-κB family [153]. Cytokine storm syndrome, which is correlated with more severeCOVID-19-related manifestations, is influenced by NF-kB. Therefore, therapeutics that inhibit the NF-κB pathway should be considered in COVID-19 therapy [154].

Additionally, NFKB may serve as a therapeutic target for tuberculosis [155]. Inhibition of NF-κB activation specifically reduces intracellular M. tuberculosis viability in human macrophages [156]. TNFalpha is involved in the development of lung cancer linked to inflammation and blocking NF-kB to stimulate apoptosis in cells of lung cancer [157][158]. Natural substances with NF-kB-suppressing activities are of significant interest in the treatment of inflammation and the prevention of lung cancer [159]. We also hypothesized that mir-124-3p, mir-26a-5p, mir-27a-3p, mir-200b-3p, mir-34a-5p, and mir-29a-3p are linked with different genes SARS CoV-2, tuberculosis and lung cancer. Among them, the overexpression of miR-124-3p resulted in the degradation of Ddx58 and a decrease in SARS-CoV-2 genome replication and effectively inhibited metastasis in non-small cell lung cancer (NSCLC) by inhibiting intracellular PI3K/AKT signaling and extracellular exosome transpor[160][161]. Upregulated miR-27a-3p expression was found in the TB patients [162]. MiR-27a-3p can suppress the NSCLC malignancy by inhibiting the MAP17 expression [163]. In COVID-19 patients, miR-200b-3p is a possible biomarker for prognosis and severity, and over-expression can enhance the Uncontrolled growth and spread of lung cancer by modulating the expression of LATS2 and SOCS6 which act as tumor inhibitors in many tumors [164][165]. In the majority of COVID-19 patients, miR-34a-5p was shown to be down-regulated in the lung biopsies and exhibited as an anti-oncogenic in LUAD [166] [167]. miR-29a-3p may serve as a potential marker for diagnosis of acute-COVID-19 disease and the overexpression of miR-29a-3p is with the increase of COVID-19 grade [168] [169]. Moreover, mir-29a-3p is a novel biomarker to diagnose active and latent pulmonary TB infection. The expression of mir-29a-3p is increased in individuals with active pulmonary tuberculosis and latent pulmonary tuberculosis. Additionally, overexpression of mir-29a-3p in LUAD cells may inhibit cell growth, migration, and infiltration [170] [171]. Overexpression of miR-148b-3p was found in untreated TB patients and it also reduced the cell migration, proliferation, invasion, and induced apoptosis of NSCLC [172] [173]. It is important to acknowledge several limitations of our study. Our analysis relied on a limited number of RNA-seq datasets (GSE171110, GSE89403, and GSE81089), which may not fully capture the diversity of genetic backgrounds and disease stages across SARS-CoV-2, tuberculosis, and lung cancer. We identified common genes and potential biomarkers, but our study did not include experimental validation of these findings. Further studies incorporating larger and more diverse datasets, along with experimental validation, are warranted to validate the clinical relevance of our findings.

## Conclusion

The current research has demonstrated a comprehensive overview of the interconnections between three disease genes within the framework of transcriptomic analysis associated with COVID-19, lung cancer, and tuberculosis. We did differential expression analysis (DEGs) on three sets of data and found the genes that overlapped. We also looked into the disease responses linked to lung cancer, tuberculosis, and SARS-CoV-2. Combining DEGs with other studies of biomolecular interactions helped us find 15 hub proteins from studies of regulatory TFs, protein-protein interactions from studies of TF-DEG interactions, and miRNAs from studies of miRNA-DEGs interactions. Each of these hub genes serves essential roles in various functional mutations in COVID-19, lung cancer, and tuberculosis, contributing to the development or progression of these disorders. In addition, these hub genes has the capacity to function as biomarkers and might possibly play a role in the development of innovative treatment strategies for the illnesses being studied. Transcription factors (TFs) regulate the speed of transcription, whereas microRNAs (miRNAs) have a vital function in overseeing gene expression and inhibiting RNA at the post-transcriptional level.. Insufficient amounts of a microRNA can lead to overactivation of genes that it controls, facilitating cancer growth and advancement. We implemented transcriptomic analysis to identify common pathways and molecular biomarkers among patients with SARS-CoV-2, lung cancer, and tuberculosis in order to gain insight into the interconnections between these three diseases. Researchers and medical specialists may also employ it as a vital strategy to discover the underlying pathophysiology and type of the specific fundamental disease in order to produce more efficacious and dependable remedies. SARS-CoV-2 specifically impacts the lower respiratory tract and can cause severe pneumonia, which significantly increases the risk of death for individuals with lung cancer. Finally, we found 35 possible therapeutic molecules for well-known differentially expressed genes (DEGs) that could be used to treat SARS-CoV-2, lung cancer, and tuberculosis in the future

## Data Availability

All data produced are available online at GSE171110, GSE89403, and GSE81089.

https://www.ncbi.nlm.nih.gov/gds

## Notes

### Competing Interest Statement

The authors have declared no competing interest.

### Funding Statement

This study did not receive any funding

